# Outcomes of laboratory-confirmed SARS-CoV-2 infection in the Omicron-driven fourth wave compared with previous waves in the Western Cape Province, South Africa

**DOI:** 10.1101/2022.01.12.22269148

**Authors:** Mary-Ann Davies, Reshma Kassanjee, Petro Rosseau, Erna Morden, Leigh Johnson, Wesley Solomon, Nei-Yuan Hsiao, Hannah Hussey, Graeme Meintjes, Masudah Paleker, Theuns Jacobs, Peter Raubenheimer, Alexa Heekes, Pierre Dane, Jamy-Lee Bam, Mariette Smith, Wolfgang Preiser, David Pienaar, Marc Mendelson, Jonathan Naude, Neshaad Schrueder, Ayanda Mnguni, Sue Le Roux, Katie Murie, Hans Prozesky, Hassan Mahomed, Liezel Rossouw, Sean Wasserman, Deborah Maughan, Linda Boloko, Barry Smith, Jantjie Taljaard, Greg Symons, Ntobeko Ntusi, Arifa Parker, Nicole Wolter, Waasila Jassat, Cheryl Cohen, Richard Lessells, Robert J Wilkinson, Juanita Arendse, Saadiq Kariem, Melvin Moodley, Krish Vallabhjee, Milani Wolmarans, Keith Cloete, Andrew Boulle, On behalf of the Western Cape and South African National Departments of Health in collaboration with the National Institute for Communicable Diseases in South Africa

**Author notes:** **Corresponding author:** Mary-Ann Davies, Affiliation: Health Impact Assessment Directorate, Western Cape Government: Health and Centre for Infectious Disease Epidemiology and Research, School of Public Health and Family Medicine, University of Cape Town, South Africa. **Email**, **Address**: University of Cape Town, Faculty of Health Sciences, Anzio Road, Observatory, 7925, CAPE TOWN, SOUTH AFRICA.

## Abstract

**Objectives:** We aimed to compare COVID-19 outcomes in the Omicron-driven fourth wave with prior waves in the Western Cape, the contribution of undiagnosed prior infection to differences in outcomes in a context of high seroprevalence due to prior infection, and whether protection against severe disease conferred by prior infection and/or vaccination was maintained.

**Methods:** In this cohort study, we included public sector patients aged ≥20 years with a laboratory confirmed COVID-19 diagnosis between 14 November-11 December 2021 (wave four) and equivalent prior wave periods. We compared the risk between waves of the following outcomes using Cox regression: death, severe hospitalization or death and any hospitalization or death (all ≤14 days after diagnosis) adjusted for age, sex, comorbidities, geography, vaccination and prior infection.

**Results:** We included 5,144 patients from wave four and 11,609 from prior waves. Risk of all outcomes was lower in wave four compared to the Delta-driven wave three (adjusted Hazard Ratio (aHR) [95% confidence interval (CI)] for death 0.27 [0.19; 0.38]. Risk reduction was lower when adjusting for vaccination and prior diagnosed infection (aHR:0.41, 95% CI: 0.29; 0.59) and reduced further when accounting for unascertained prior infections (aHR: 0.72). Vaccine protection was maintained in wave four (aHR for outcome of death: 0.24; 95% CI: 0.10; 0.58).

**Conclusions:** In the Omicron-driven wave, severe COVID-19 outcomes were reduced mostly due to protection conferred by prior infection and/or vaccination, but intrinsically reduced virulence may account for an approximately 25% reduced risk of severe hospitalization or death compared to Delta.

## Background

Following the identification and early spread of the Omicron (B.1.529) SARS-CoV-2 variant of concern (VOC) during November 2021, South Africa observed the steepest surge in COVID-19 cases to date (1, 2). With more than 50 mutations across its genome, in vitro, ex vivo and modeling studies have uncovered potential changes in the biology of Omicron compared to previous VOCs, such as tropism, immune escape and improved transmissibility (3-8).

South Africa had previously experienced three COVID-19 waves related to different SARS-CoV-2 variants (ancestral strain, Beta and Delta respectively), each more clinically severe than the previous one with substantial mortality (9, 10). These waves have resulted in high seroprevalence of ∼70% from prior infection, especially in poorer communities where social distancing is challenging (11, 12). While such high seroprevalence came at the cost of exceptionally high mortality during the first wave of COVID-19, these areas were relatively protected from both infections and severe disease in subsequent waves (13). For example, in the large urban township of Khayelitsha, the poorest subdistrict in Cape Town, anti-nucleocapsid antibody seroprevalence resulting from prior infection was 45% by the end of wave one (11), increasing to >70% by the end of wave three (Hsiao, NY., personal communication). Unlike the relatively small and slow waves two and three, evidence of immune escape from infection with Omicron is clearly illustrated by the steep rise in cases in Khayelitsha in wave four, similar to that of wave one, in comparison to waves two and three (Figure 1).

**Figure 1:**
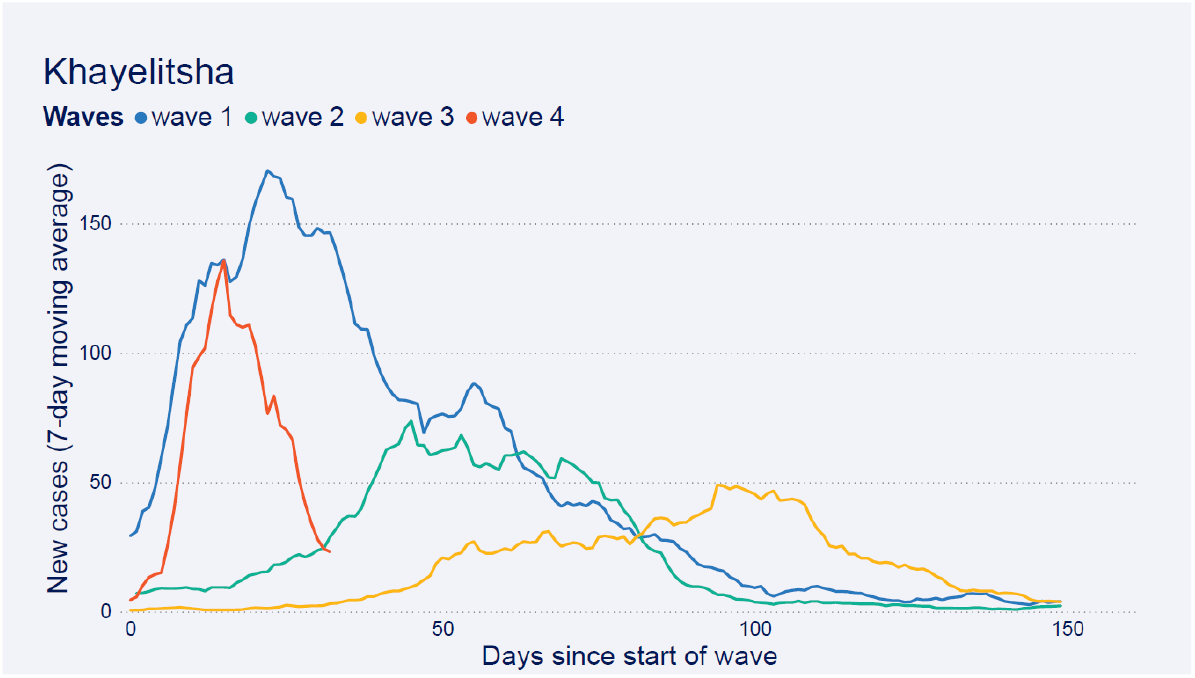
Daily new cases (7 day moving average) by days since start of each wave in Khayelitsha sub-district, Cape Town, South Africa.

While there is emerging biological evidence of possible lower virulence of Omicron compared to previous variants due to modified cell entry mechanisms and preferential replication in bronchi rather than the lung parenchyma (6, 7, 14, 15), even a virus resulting in similar or lower clinical severity to earlier variants could overwhelm health services if protection conferred by prior COVID-19 infection and/or SARS-CoV-2 vaccination against severe disease is reduced. While there are now several reports from South Africa and other countries of reduced risk of severe disease in the fourth wave and in patients infected with Omicron compared to Delta (16-21) in the context of a high seroprevalence setting such as South Africa, with moderate SARS-CoV-2 vaccination coverage (39% and 46% of adults fully vaccinated in South Africa and the Western Cape respectively by end December 2021) (22), it is important to establish whether protection against severe disease conferred by prior infection and/or vaccination is maintained against Omicron. Furthermore, to what extent does such protection account for milder clinical presentation of Omicron cases versus inherent differences in virulence of Omicron itself compared to previous variants? Such comparisons should not be limited to Delta, which was itself more severe than previous variants (23-26), and should fully account for the increased proportion of reinfections in an immune escape variant such as Omicron compared to other variants (18, 27). While Omicron’s immune evasion allows for vaccine breakthroughs and reinfections, if protection against severe disease conferred by prior infection and/or vaccination is maintained, population data should reflect milder clinical illness due to a higher proportion of Omicron cases with reinfections/breakthroughs compared to cases of variants without escape from immunity against infection (18, 27).

We compared outcomes of laboratory-confirmed SARS-CoV-2 infections across four successive waves in those aged ≥20 years using public sector services in the Western Cape Province, South Africa, accounting for prior infection and vaccination. We also assessed whether protection against severe disease conferred by prior diagnosed infections and/or vaccination was maintained in those infected during the Omicron-driven fourth wave, and examined the extent to which undiagnosed prior infection may account for observed reductions in clinical severity during the Omicron wave.

## Methods

### Study design

We conducted a cohort study using de-identified data from the Western Cape Provincial Health Data Centre (WCPHDC) of public sector patients aged ≥20 years with a laboratory confirmed COVID-19 diagnosis (positive SARS-CoV-2 PCR or antigen test). For this analysis, each wave was deemed to commence on the date when COVID-19 hospital admissions in public sector patients showed a sustained >10% week-on-week increase. We included cases diagnosed from seven days before the wave start (to account for the lag between infection/first symptoms and hospitalization) and for the following four weeks, to allow for at least 2 weeks of follow up in the most recently diagnosed patients in wave four. We thus included data for the 4^th^ wave on cases from 14 November to 11 December 2021, with follow-up through to 26 December 2021, which corresponds to the period when omicron rapidly became the dominant variant in the province accounting for nearly 100% of sequenced cases.(2) Database closure was ten days later to allow for death reporting delays. For the first wave, since there were no prior admissions, we selected a period where case incidence was the same as that at the start of wave 4.

The study was approved by the University of Cape Town and Stellenbosch University Health Research Ethics Committees and Western Cape Government: Health. Individual informed consent requirement was waived for this secondary analysis of de-identified data.

### Study population and data sources

The Western Cape has nearly 7 million inhabitants, of whom approximately 75% use public sector health services (28). The WCPHDC has been described in detail (10, 29). Briefly, WCPHDC consolidates administrative, laboratory, and pharmacy data from routine electronic clinical information systems used in all public sector health facilities with linkage through a unique identifier. Multiple data sources are triangulated to enumerate health conditions such as diabetes mellitus (“diabetes”), hypertension, tuberculosis and HIV-1. Hospitalizations (private and public) and reported deaths with a positive SARS-CoV-2 laboratory test are recorded and reviewed daily. For patients with recorded South African national identity numbers, data are linked to the South African vital registry to identify deaths not recorded in the WCPHDC. SARS-CoV-2 vaccination commenced on 17 February 2021 for health workers and 17 May 2021 for the general population in age cohorts, starting with those aged ≥60 years and progressively expanding to younger ages. By 20 October 2021, vaccination was available to all individuals aged ≥12 years. All SARS-CoV-2 vaccinations administered in the country are recorded on the Electronic Vaccine Data System (EVDS). Vaccination data (dates and types of all SARS-CoV-2 vaccines given at any facility in the province) were obtained using the South African national identifier to link WPHDC data to EVDS data for a specific individual.

### Statistical analysis

We assessed three outcomes: (i) death, (ii) severe hospitalization or death and (iii) any hospitalization or death. We only included outcomes within 14 days of COVID-19 diagnosis to allow for comparable ascertainment across all wave periods. Hospitalization included admission within 14 days before or after a COVID-19 diagnosis except for admissions to long term psychiatric or rehabilitation facilities where the COVID-19 diagnosis was likely to be incidental. Severe hospitalization was defined as admission to intensive care or mechanical ventilation or oral/intravenous steroid prescription. Deaths within 14 days of COVID-19 diagnosis were included unless a clear non-COVID-19 cause of death was recorded.

We used Cox regression adjusted for age, sex, geographic location, comorbidities, SARS-CoV-2 vaccination and prior diagnosed infection to assess differences in COVID-19 outcomes between waves. Vaccination at the time of COVID-19 diagnosis was defined as “fully” (≥28 days post single dose vaccination with Janssen/Johnson & Johnson [Ad26.COV2.S], or ≥14 days post second dose of Pfizer–BioNTech [BNT162b2]), or “partially” (≥21 days after single dose Ad26.COV2.S vaccine or first BNT162b2 dose until meeting criteria for fully vaccinated). Additional booster doses were not considered as these were not widely available except to health care workers and the severely immune compromised. Prior diagnosed infection was defined as ≥1 laboratory confirmed SARS-CoV-2 diagnosis ≥90 days previously without an intervening positive test. We used the approach described by Ferguson et al. (18) to assess the extent to which reductions in disease severity during the Omicron period may be attenuated by more unascertained prior infections in those with Omicron compared to patients infected with previous variants. As a base scenario we assumed that immunity conferred by prior infection reduces risk of death, severe hospitalization or death and hospitalization or death by 80%, 80% and 70% respectively, and that only 15% of prior diagnosed infections were ascertained based on seroprevalence and excess death data (12, 30). We then calculated a corrected hazard ratio for wave four vs. wave three using the formula:

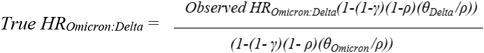

where γ is the relative risk of the outcome in those with vs. without prior infection, ρ is the proportion of reinfections detected and θ is the observed proportion of reinfections with Delta and Omicron respectively (18). The proportional-hazard assumption (assessed with Schoenfeld residuals) (31) was violated for the effect of wave on the outcome of any hospitalization or death only, as the hazards converged over time. We therefore also show the results of logistic regression for all of the outcomes which were very similar.

In addition, to assess whether protection conferred by prior infection and vaccination was similar in the Omicron and Delta periods, we compared the association between these variables and the three severe COVID-19 outcomes separately for just the wave three and wave four periods. The wave three period was cases diagnosed between 1 September and 15 October 2021 as too few people had been vaccinated during early wave three (26 May to 23 June 2021) since vaccination only commenced for those 60 years and older on 17 May 2021. All analyses were conducted using Stata 17.1.

## Results

We included 5,144 patients diagnosed in wave four and 4,403, 3,902 and 3,304 patients from waves three, two and one respectively (Table 1). There was a greater proportion of patients aged 20-39 years in wave four (64%) compared to waves two (49%) and three (44%). The prevalence of comorbidities was mostly similar across waves, except for HIV-1 which had highest prevalence in wave one, decreased prevalence in waves two and three, but increased prevalence in wave four (Table 1). The proportion with prior diagnosed infection was substantially higher in wave four (11%) compared to waves three (3.2%) or two (1.9%). In wave four, 38% and 5% of all COVID-19 cases were fully or partially vaccinated respectively.

**Table 1:**
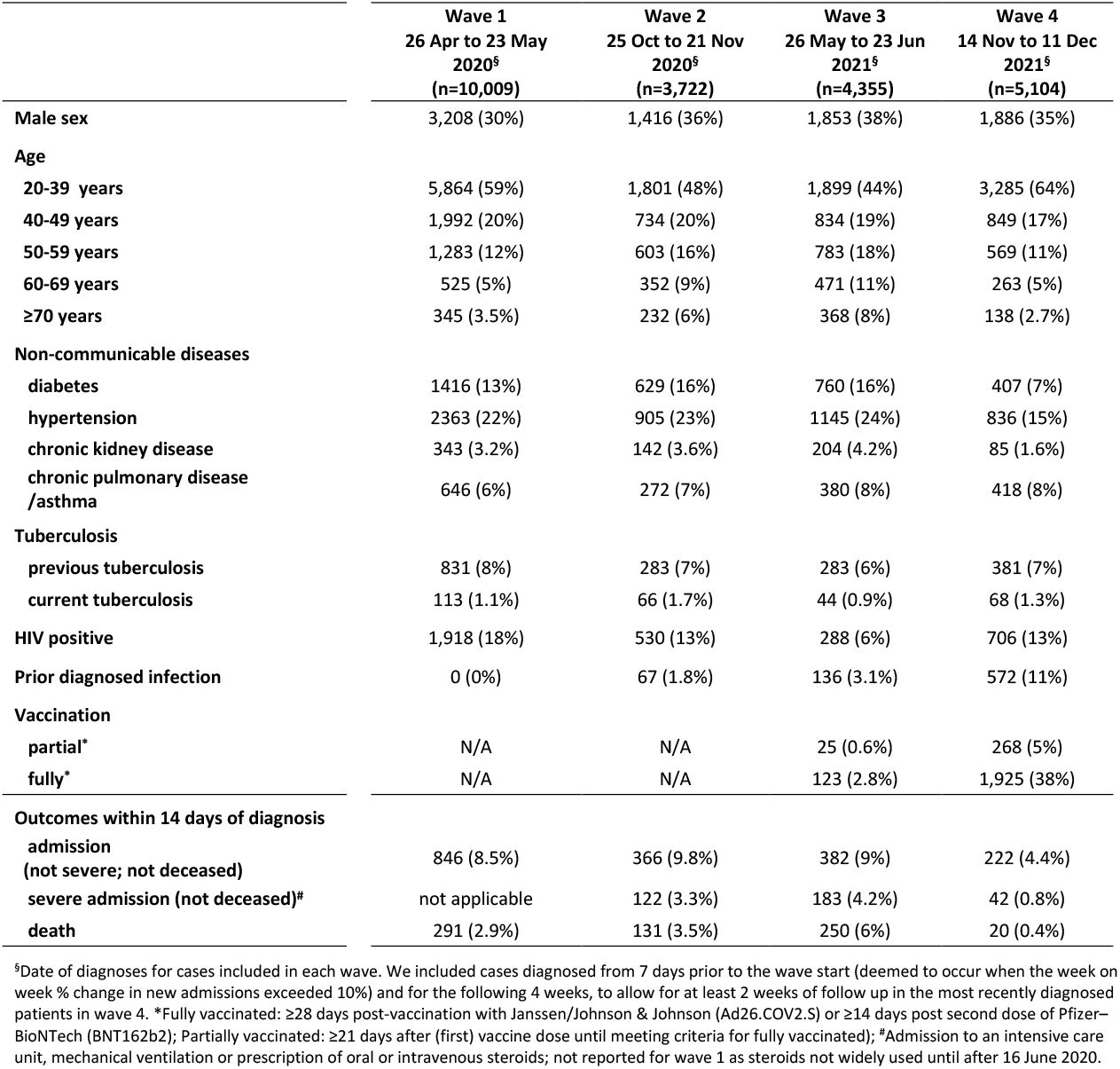
Characteristics and outcomes of COVID-19 cases included from each of the four waves in the Western Cape Province, South Africa.

### Comparison of outcomes across waves

Overall, 8.0% (409/5,144) of cases were hospitalized or died within 14 days of diagnosis in wave four compared to 16.5% (1918/11,609) across the previous three waves (Table 1). After adjusting for age, sex, comorbidities and subdistrict there was a substantially reduced hazard of death in wave four compared to wave three (adjusted Hazard Ratio [aHR] 0.27; 95% Confidence Interval: 0.19; 0.38) (Table 2). The extent of reduction was attenuated (0.41; 95% CI 0.29; 0.59) when additionally considering prior diagnosed infections and vaccination. Vaccination was strongly protective (aHR for fully vs. not vaccinated 0.20; 95% CI 0.09; 0.43). Wave four was also associated with lower risk of death than waves one and two, which in turn were less severe than wave three (aHR [95% CI] for waves one and two vs wave three were 0.55 [0.40; 0.74] and 0.60 [0.48; 0.74] respectively). The pattern of reduced severity in wave four compared to previous waves was similar for the outcome of severe hospitalization or death, but for the least specific outcome (i.e. any hospitalization or death) the risk reduction in wave four vs. three was smaller. For all outcomes, the reduced risk of the outcome attenuated with adjustment for prior diagnosed infection and vaccination. For example, for the outcome of any hospitalization or death, the risk was lower for wave four vs wave three (aHR 0.72; 95% CI: 0.63; 0.82) and wave two, but greater than wave one (aHR [95%CI] for waves two and one vs wave three: 0.88 [0.80; 0.96] and 0.57 [0.50; 0.66] respectively). Both prior diagnosed COVID-19 (aHR 0.28; 95% CI 0.19; 0.40) and SARS-CoV-2 vaccination were protective (aHR 0.42; 95% CI 0.34; 0.52). Results were very similar when using logistic regression (Supplementary Table 1).

**Table 2:** Associations between different waves and severe COVID-19 outcomes adjusted for patient characteristics, sub-district, vaccination, prior diagnosed infection and unascertained prior infections using Cox regression.

|  | Outcome = death<br>not adjusted for<br>vaccination and prior<br>infection |  | Outcome = death<br>adjusted for vaccination<br>and prior infection |  | Outcome = severe<br>hospitalization#/death<br>not adjusted for<br>vaccination or prior<br>diagnosed infection |  | Outcome = severe<br>hospitalization#/death<br>adjusted for vaccination<br>or<br>prior diagnosed infection |  | Outcome =<br>hospitalization/death<br>not adjusted for<br>vaccination and prior<br>infection |  | Outcome =<br>hospitalization/death<br>adjusted for vaccination<br>and prior infection |  |
| --- | --- | --- | --- | --- | --- | --- | --- | --- | --- | --- | --- | --- |
|  | Adjusted<sup>^</sup><br>HR | 95% CI | Adjusted<br>HR | 95% CI | Adjusted<sup>^</sup><br>HR | 95% CI | Adjusted<br>HR | 95% CI | Adjusted<sup>^</sup><br>HR | 95% CI | Adjusted<br>HR | 95% CI |
| Male sex (vs. female) | 1.44 | 1.20; 1.73 | 1.45 | 1.21; 1.74 | 1.37 | 1.19; 1.56 | 1.37 | 1.19; 1.57 | 1.15 | 1.06; 1.24 | 1.14 | 1.06; 1.23 |
| Age (vs. 20-29 years) |  |  |  |  |  |  |  |  |  |  |  |  |
| 40-49 years | 3.07 | 1.96; 4.80 | 3.18 | 2.03; 4.98 | 2.32 | 1.74; 3.10 | 2.42 | 1.82; 3.24 | 1.14 | 1.00; 1.29 | 1.17 | 1.03; 1.33 |
| 50-59 years | 7.35 | 4.88; 11.06 | 7.64 | 5.07; 11.52 | 4.74 | 3.66; 6.12 | 4.96 | 3.82; 6.42 | 1.90 | 1.68; 2.15 | 1.96 | 1.73; 2.21 |
| 60-69 years | 17.17 | 11.31; 26.05 | 18.11 | 11.92; 27.52 | 8.51 | 6.52; 11.11 | 8.85 | 6.77; 11.58 | 2.94 | 2.57; 3.36 | 3.01 | 2.63; 3.45 |
| ≥70 years | 30.94 | 20.10; 47.62 | 34.22 | 22.22; 52.68 | 13.56 | 10.30; 17.85 | 14.39 | 10.91; 18.98 | 4.25 | 3.70; 4.89 | 4.40 | 3.83; 5.06 |
| Comorbidities (vs. comorbidity absent) |  |  |  |  |  |  |  |  |  |  |  |  |
| diabetes | 1.97 | 1.60; 2.41 | 1.95 | 1.58; 2.39 | 1.76 | 1.51; 2.05 | 1.77 | 1.51; 2.06 | 2.22 | 2.02; 2.44 | 2.26 | 2.06; 2.48 |
| hypertension | 1.06 | 0.87; 1.30 | 1.05 | 0.86; 1.28 | 1.01 | 0.87; 1.18 | 1.00 | 0.86; 1.17 | 1.11 | 1.01; 1.21 | 1.10 | 1.0; 1.20 |
| chronic kidney disease | 2.04 | 1.60; 2.60 | 2.07 | 1.63; 2.63 | 1.73 | 1.43; 2.09 | 1.73 | 1.43; 2.09 | 1.71 | 1.52; 1.92 | 1.70 | 1.52; 1.91 |
| chronic pulmonary disease / asthma | 1.24 | 0.97; 1.59 | 1.24 | 0.97; 1.59 | 1.44 | 1.20; 1.73 | 1.46 | 1.21; 1.75 | 1.24 | 1.11; 1.39 | 1.27 | 1.13; 1.42 |
| previous tuberculosis | 1.70 | 1.24; 2.33 | 1.68 | 1.22; 2.29 | 1.39 | 1.09; 1.78 | 1.38 | 1.08; 1.76 | 1.17 | 1.02; 1.34 | 1.16 | 1.01; 1.33 |
| current tuberculosis | 2.24 | 1.19; 4.21 | 2.09 | 1.11; 3.91 | 3.13 | 2.10; 4.67 | 2.99 | 2.01; 4.47 | 3.25 | 2.63; 4.03 | 3.24 | 2.63; 3.99 |
| HIV | 1.88 | 1.36; 2.58 | 1.92 | 1.40; 2.64 | 1.49 | 1.16; 1.92 | 1.53 | 1.19; 1.96 | 1.58 | 1.40; 1.78 | 1.61 | 1.43; 1.81 |
| Prior diagnosed infection (vs. None) |  |  |  |  |  |  |  |  |  |  |  |  |
| Yes (vs none) |  |  | 1.10 | 0.63; 1.92 |  |  | 0.60 | 0.37; 0.98 |  |  | 0.28 | 0.19; 0.40 |
| Vaccination (vs. None) |  |  |  |  |  |  |  |  |  |  |  |  |
| partial<sup>*</sup> |  |  | 1.19 | 0.66; 2.15 |  |  | 1.26 | 0.81; 1.95 |  |  | 0.89 | 0.66; 1.20 |
| full<sup>*</sup> |  |  | 0.20 | 0.09; 0.43 |  |  | 0.23 | 0.13; 0.39 |  |  | 0.42 | 0.34; 0.52 |
| "Wave period" |  |  |  |  |  |  |  |  |  |  |  |  |
| early wave 1 | 0.59 | 0.43; 0.81 | 0.55 | 0.40; 0.77 |  |  |  |  | 0.60 | 0.53; 0.69 | 0.57 | 0.50; 0.66 |
| early wave 2 | 0.64 | 0.52; 0.80 | 0.60 | 0.48; 0.74 | 0.74 | 0.64; 0.85 | 0.71 | 0.61; 0.83 | 0.91 | 0.83; 0.99 | 0.88 | 0.80; 0.96 |
| early wave 3 | Ref | Ref |  |  | Ref | Ref | Ref | Ref | Ref | Ref | Ref | Ref |
| early wave 4 | 0.27 | 0.19; 0.38 | 0.41 | 0.29; 0.59 | 0.28 | 0.22; 0.36 | 0.43 | 0.33; 0.55 | 0.51 | 0.46; 0.58 | 0.72 | 0.63; 0.82 |
| early wave 4 accounting for<br>undiagnosed prior infections<sup>§</sup> |  |  | 0.72 |  |  |  | 0.75 |  |  |  | 1.14 |  |

\*Fully vaccinated: ≥28 days post-vaccination with Janssen/Johnson & Johnson (Ad26.COV2.S) or ≥14 days post second dose of Pfizer–BioNTech (BNT162b2); Partially vaccinated: ≥21 days after (first) vaccine dose until meeting criteria for fully vaccinated); <sup>^</sup>Admission to an intensive care unit, mechanical ventilation or prescription of oral or intravenous steroids; not reported for wave 1 as steroids not widely used until after 16 June 2020. <sup>^</sup>Adjusted for all variables shown in the table as well as subdistrict/district, but not for vaccination or prior diagnosed infection; <sup>§</sup>We calculated the "true hazard ratio" for severity of wave 4 vs wave 3 using the approach described by Ferguson et al. (18), assuming prior infection reduces risk of death, severe admission or death and admission or death by 80%, 80% and 70% respectively, and that 15% of prior diagnosed infections were ascertained. aHR = adjusted Hazard Ratio; CI = Confidence Interval

After considering the possible effect of protection against severe outcomes conferred by unascertained prior infection, the reduced risk of severe disease in wave four vs wave three remained but was substantially attenuated with aHR of 0.72 for death and 0.75 for severe hospitalization or death respectively, and the risk of any COVID-19 hospitalization or death was similar or higher in wave four and wave 3 (aHR: 1.14). Results were sensitive to the extent of protection assumed to be provided from prior infection and the proportion of prior infections assumed to be ascertained (Supplementary Table). For example, there was no difference in risk of severe hospitalization or death in wave three vs. wave four if the assumed proportion of prior infections detected was reduced from 15% to 12%.

### Protection from vaccination and prior infection in waves three and four

Amongst COVID-19 cases, protection by vaccination against all outcomes was similar in wave four compared to the prior wave (Figure 2). For example, the aHR (95% CI) for protection against death from full vaccination was 0.35 (0.22; 0.54) in late wave three and 0.24 (0.10; 0.58) in wave four. Similarly, protection conferred by prior diagnosed infection against hospitalization or death was maintained with aHR (95%CI) of 0.32 (0.20; 0.52) in late wave three and 0.13 (0.06; 0.27) during wave four. Protection conferred by prior infection against other severe COVID-19 outcomes was difficult to assess due to very small numbers of patients with prior diagnoses experiencing these outcomes.

**Figure 2:**
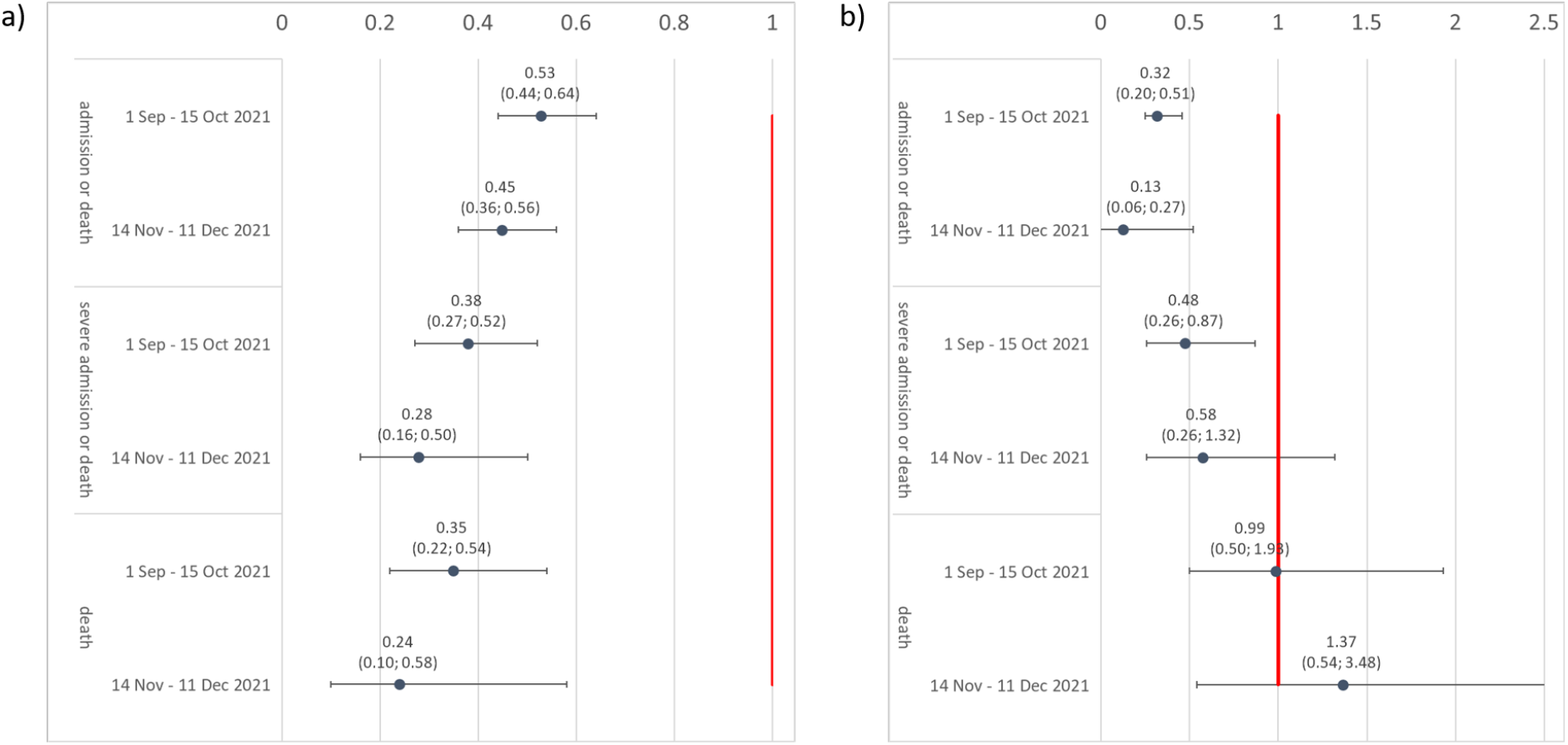
Adjusted hazard ratio for associations between a) vaccination and b) prior diagnosed infection and different severe COVID-19 outcomes adjusted for patient characteristics, subdistrict, vaccination, and prior diagnosed infection using Cox regression.

## Discussion

In this study of Western Cape public sector laboratory confirmed COVID-19 cases, we found a substantial reduction in all severe COVID-19 outcomes in wave four compared to previous waves with 28% (95% CI: 18%; 37%) reduced risk of any hospitalization or death and 59% (95% CI: 41%; 71%) reduced risk of death only after adjusting for vaccination. While some of this reduction is likely due to retained protection conferred by prior SARS-CoV-2 infection against severe disease in Omicron-infected patients, even when accounting for protection conferred by prior unascertained infections in patients with Omicron, we found approximately 25% reduction in severe hospitalization or death in wave four vs wave three.

Our findings concur with current literature reporting less severe disease associated with Omicron vs. other variants of concern and previous waves (16-20, 32). For example, cases with presumed Omicron infection (identified with S-gene target failure on PCR) were less likely to be admitted to hospital and experience severe outcomes compared to those with delta infections (18, 19, 21) and fewer admissions with less severe outcomes among those hospitalized have been reported in the fourth wave compared to previous waves in South Africa (16, 17). To our knowledge, ours is the first study from a setting of high prior seroprevalence to demonstrate less severe disease in wave four after adjusting for both vaccination and prior diagnosed infection, and to assess the contribution to protection of a greater proportion of unascertained re-infections among Omicron cases compared to other variants. Reassuringly, protection against severe disease conferred by prior infection and vaccination were similar in wave four and wave three. Nonetheless, the fact that even after this protection was considered there was likely reduction of the most severe outcomes in wave four indicates possible reduction in virulence of Omicron. This is supported by laboratory data from several studies demonstrating possible mechanisms of reduced virulence (6-8, 14).

The finding of increased prevalence of HIV-1 in wave four compared to the previous two waves illustrates the importance of adjusting for prior unascertained infections. Poorer urban communities tend to have higher HIV-1 prevalence and so the pattern of decreasing HIV-1 prevalence in the second and third waves, with increased prevalence among fourth wave cases, is unlikely to be due to HIV-1 itself, but because HIV-1 infection is a proxy for living in a community like Khayelitsha subdistrict, with high SARS-CoV-2 seroprevalence by the end of the first wave, which protected against infection in the two subsequent waves, but less so in wave four.

For the less severe outcome of any hospitalization or death, while risk in wave four was less than wave three, after adjusting for vaccination and prior diagnosed infections it was similar to that of wave one. This is notable as wave three was driven by the Delta variant which has been shown to cause more severe disease than ancestral strains (23-26). Hospitalization risk may appear similar to or higher in wave four compared to earlier waves which were due to less transmissible variants simply due to a higher prevalence of cases during the wave surge together with more widespread testing of asymptomatic hospitalized patients than in previous waves. Notwithstanding, the similarity in risk of admission suggests that in the absence of immunity, Omicron could be as severe as the ancestral strain. Irrespective of virulence and disease severity, the sheer number of admissions in patients during an Omicron wave warrants specific planning around appropriate infection prevention and control measures within hospitals while minimizing adverse impacts on health services for other conditions.

Strengths of our study include complete ascertainment of hospitalizations and deaths in all laboratory confirmed COVID-19 cases across a public sector health service and ability to robustly adjust for comorbidities as well as SARS-CoV-2 vaccination. Our analysis also has several limitations. First, we compared outcomes across waves as a proxy for the variant that dominated in each wave, and not in patients with genomically confirmed variants. Second, while health service pressures which impact disease outcomes are likely to be more similar at “equivalent wave periods”, identifying such “equivalent periods” across waves can be challenging. However, results were similar when using slightly different wave periods. Third, adjustment for comorbidities was limited to those algorithmically identified in the WCPHDC and does not include undiagnosed comorbidities and other important risk factors for poor COVID-19 outcomes such as obesity. Fourth, prior diagnosed infections substantially under-ascertain all prior infections, and while we addressed this by determining the likely impact of undiagnosed infections in wave four, this is based on assumptions. Fifth, we could not distinguish between admissions and deaths where the diagnosis of COVID-19 may have been incidental or contributory rather than causal. However, our main analysis focused on mortality and we found stronger protection of both wave four and vaccination against the more severe outcomes (severe hospitalization and/or death), suggesting that results are robust despite misclassification of admissions with incidental COVID-19. Nonetheless, a different clinical profile of hospitalized and deceased patients with COVID-19 has been reported, with less COVID-19 pneumonia and a greater proportion of patients with severe comorbidities where COVID-19 may be contributory, but not causing typical respiratory presentation (17). We may therefore be underestimating the reduction in risk of hospitalization and death due to COVID-19 pneumonia specifically in the Omicron wave. Sixth, we excluded children as the effects of Omicron on disease severity may differ in children compared to adults, the role of COVID-19 as a cause of pathology in a child admitted with respiratory illness with several different viruses present is unclear, and because we could not assess differences in the more severe outcomes between waves in children as these were so uncommon. Finally, outcomes were limited to 14 days post diagnosis to allow for equivalent follow-up in the most recent vs. previous waves and so we could not compare outcomes beyond 14 days. However, in previous waves only 2.9% of hospitalizations occurred beyond this period and only 2.6% of deaths had not been hospitalized or deceased within 2 weeks of diagnosis.

In conclusion, we found substantially reduced disease severity amongst diagnosed COVID-19 cases in the Omicron-driven fourth wave compared to previous waves. While this could mostly be due to retained protection against severe outcomes conferred by prior infection and vaccination, our data suggests that severe outcomes could be reduced by approximately 25% due to intrinsically reduced virulence of Omicron.

## Data Availability

All data produced in the present study are available upon reasonable request to the authors.

## Acknowledgements

We would like to acknowledge all patients in the Western Cape and to thank the Western Cape Department of Health Provincial Health Data Centre, the Western Cape Department of Health COVID-19 Outbreak Response Team, the Western Cape Communicable Disease Control sub-directorate and Western Cape health care workers involved in the COVID-19 response for their contributions to this report. We acknowledge funding for the Western Cape Provincial Health Data Centre from the Western Cape Department of Health, the US National Institutes for Health (R01 HD080465, U01 AI069924), the Bill and Melinda Gates Foundation (1164272, 119327), the United States Agency for International Development (72067418CA00023), the European Union (101045989), the Wellcome Trust (203135/Z/16/Z, 222574) and the Medical Research Council of South Africa. RJW receives support from the Francis Crick Institute which is funded by Wellcome (FC0010218), MRC (UK) (FC0010218) and Cancer Research UK (FC0010218). He also receives support from Wellcome (203135, 222574) and the Medical Research Council of South Africa.

---

**Supplementary Table 1:**
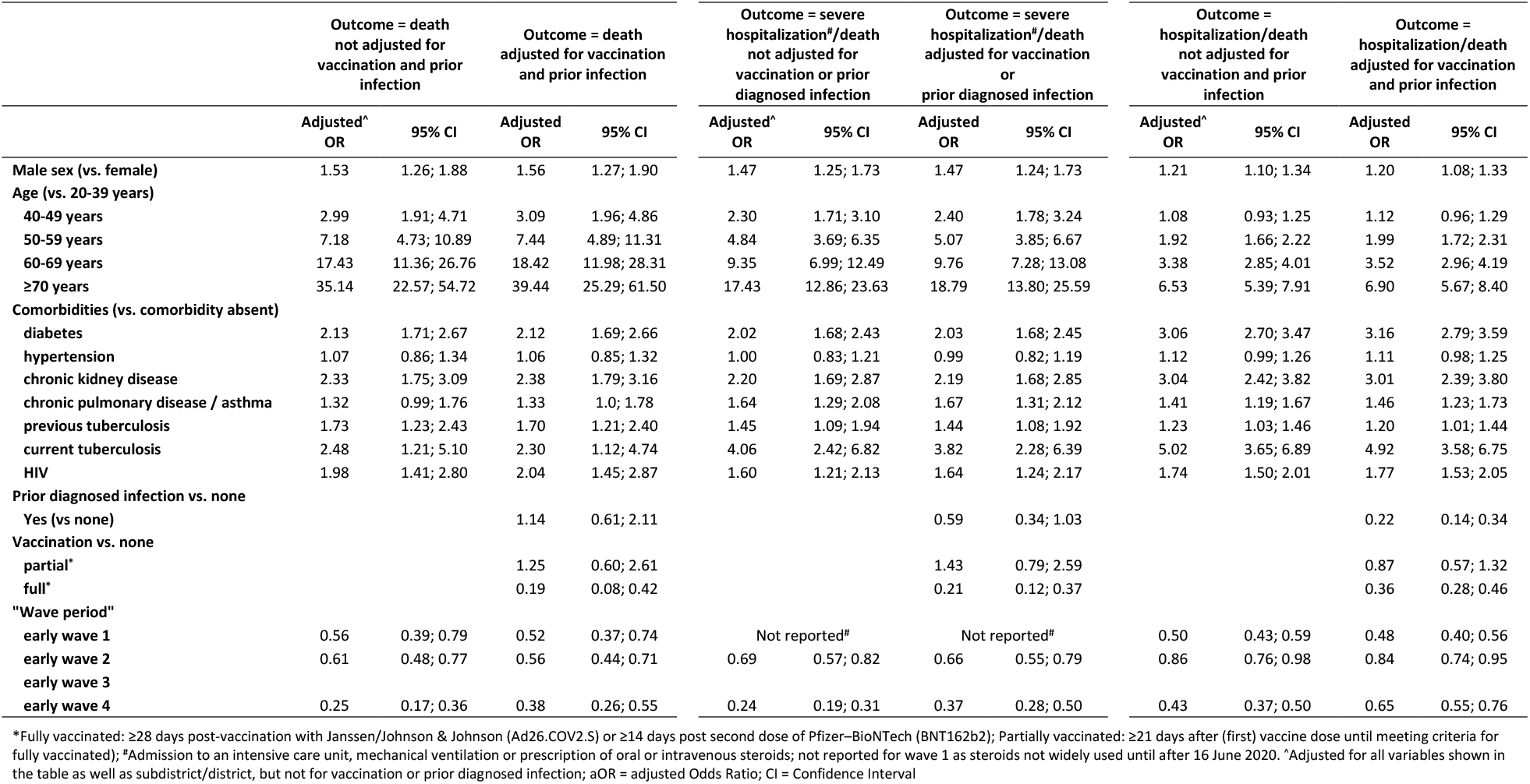
Associations between different waves and severe COVID-19 outcomes adjusted for patient characteristics, sub-district, vaccination and prior diagnosed infection using logistic regression.

**Supplementary Table 2:**
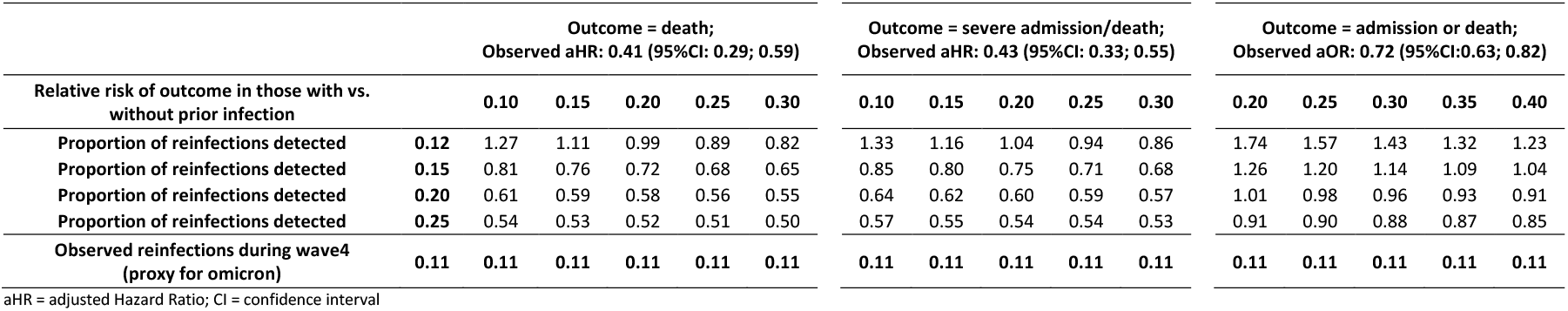
Range of possible values for the true hazard ratio for the association between wave four (vs. wave three) and each COVID-19 outcome for different assumptions of the protection that prior infection provides against the outcome and the proportion of reinfections detected among cases diagnosed during the fourth wave.

## Notes

### Competing Interest Statement

The authors have declared no competing interest.

### Author Declarations

Human Research Ethics Committee, University of Cape Town Faculty of Health Sciences, South Africa

